# Association of rs12722 COL5A1 with Pulmonary Tuberculosis infection: a preliminary case-control study in a Kazakhstani population

**DOI:** 10.1101/19008995

**Authors:** Sanzhar Zhetkenev, Ayauly Khassan, Armanay Khamzina, Alpamys Issanov, Byron Crape, Ainur Akilzhanova, Ulan Kozhamkulov, Dauren Yerezhepov, Chee Kai Chan

## Abstract

**Background:** Lung cavitation is the classic hallmark of TB, which facilitates the disease development and transmission. It involves the degradation of lung parenchyma which is mainly made up of collagen fibers by metalloproteinases (MMPs) produced by activated monocyte-derived cells, neutrophils and stromal cells.

**Objective:** The following population-based preliminary case-control study of adults with TB and controls without TB will check the possible association between rs1800012 in COL1A1, rs1272222 in COL5A1 genes to human TB susceptibility in Kazakhstan.

**Methods:** In this case-control study including 165 samples we examined the associations between TB disease status and demographic variables along with single nucleotide polymorphisms related to COLA1 and COL5A1. The unadjusted χ2 and adjusted logistic regression was performed to find out relationships between SNPS and other predictors.

**Results:** Preliminary findings suggest that there is a statistically significant association of age (OR=0.44, 95% CI:0.21-0.92, p-value=0.03), social status (OR=0.42, 95% CI:0.201 −0.87, p-value=0.020), HIV status(OR=6.9, 95% CI:1.86 - 25.6, p-value=0.004) and heterozygous rs12722 SNP (OR=2.45, 95% CI:1.16 −5.16 p-value=0.019) polymorphism of COL5A1 gene with TB susceptibility.

**Conclusion:** The association of collagen genes with TB pathogenesis indicates that anti TB programs should develop new drug regimens that include MMP inhibitors. Therapeutic targeting of MMPs will prevent extracellular matrix degradation and granuloma maturation.

## INTRODUCTION

Tuberculosis (TB) disease remains a critical health threat to the world, with 10 million infected people and 1.6 million deaths in 2017 (WHO global tuberculosis report, 2018). Despite the steady decrease in its morbidity and mortality over the last decades, the total eradication of the disease is still challenging. This is in part due to the emergence of drug-resistant strains of *M. tuberculosis*, making available TB drugs ineffective (Klopper et al., 2013).

Currently worldwide the multidrug-resistant TB rate (MDR-TB) is on the rise and is a major public health concern. The World Health Organization (WHO) estimates that there will be 558,000 new cases of TB which are resistant to rifampicin, the most effective first-line TB drug (WHO, 2019). Kazakhstan is currently among the 30 highest MDR-TB burden countries, making Kazakhtani population especially vulnerable to TB infection (“Countries with TB - High & low burden TB countries”, 2019).

Tuberculosis is transmitted via aerosol. A person becomes infected after inhalation of droplets containing *Mycobacterium tuberculosi*s (Kaufmann, 2001). The tubercle bacilli are internalized through phagocytosis by alveolar macrophages. The activated macrophages transfer the phagocytosed *M. tuberculosis* into the lysosome for destruction, but some mycobacteria are able to survive, escape lysosomal degradation and replicate inside the macrophage (Pieters, 2008; Russel, 2001). The infected macrophages can either remain in the lungs or spread to other organs in the body. Localized inflammatory response is initiated, activating the host’s immune defense system (Ramakrishnan, 2012). The immune cells, in particular macrophages and T lymphocytes, surround the *M. tuberculosis* infected macrophages, forming structured clusters called granulomas (Silva Miranda, Breiman, Allain, Deknuydt & Altare, 2012). The activated macrophages present mycobacterial antigens to the surrounding T lymphocytes within the granuloma (Chan et al., 2004). The activated T cells secrete cytokines and chemokines, ensuring the further recruitment of other immune cells to the infection site. *M. tuberculosis* can persist for a long time in a “dormant” state within the granuloma, resulting in latent infection (Karakousis et al., 2004; Dannenberg, 2003). If the immune system of the infected person is effective, there may be no adverse effect on the host’s health (Saunders et al., 2007). Activated monocyte-derived cells, neutrophils and stromal cells however produce not only various types of cytokines, but also proteases such as matrix metalloproteinases (MMPs) and cathepsin (Manka et al., 2012). The abundant production of proteases can lead to tissue damage which leads to lung cavitation.

Lung cavitation is the classic hallmark of TB disease and is the site of high mycobacterial burden, which facilitates disease development and transmission. It is associated with antibiotic resistance (Kempker et al., 2012) and treatment failure (Chatterjee et al., 2010). The formation of cavities involves the destruction of lung parenchyma, which is mainly composed of collagen fibers. Collagen type I, III and IV are the major structural components of the lung (Bateman et al.,1981). Collagen fibrils are highly resistant to enzymatic activity and can be degraded by specific proteases such as metalloproteinases (MMPs). Immune reactions initiated to eliminate foreign particles induce destruction in the lung (Ong et al., 2013).

Collagen type I protein is one of the most abundant proteins of bone matrix, which is encoded by C*OL1A1* and *COL1A2*. Mutations in the Col1A1 gene can cause osteogenesis imperfecta, a disease characterised by moderate to severe bone fragility (Singth et. al., 2011). The allelic variants of Col1A1 has been associated with bone mineral density (BMD) and risk of fracture. The most studied polymorphism of Col1A1 is rs1800012, which is located at the binding site for the transcription factor Sp1 in the first intron of Col1A1 (Grant et al., 1996). It has been associated with low BMD and increased risk of fractures because it influences DNA binding, gene transcription, protein production and bone mineralization (Jin et al., 2011; Grant et al., 1996). The TT and CC genotypes of rs1800012 have different affinities for Sp1, resulting in allele-specific transcription. Thus, T and C alleles are spliced differently within the nucleus affecting bone mineralization (Mann et al., 2001). The difference in gene variants of rs180012 can also affect its degradation by MMPs, influencing TB infection.

Collagen type V is the main structural component of tendons and ligaments (Birk 2001). Type V collagen is not a major player in the collagen hierarchy, but it has an important regulatory function. The assembly (fibrillogenesis), diameter and density of the collagen fibril within connective tissues are regulated by type V collagen through its interactions with type I collagen (Sun M et.al 2011; Wenstrup et.al 2011). The type V collagen consist of two α1(V) and one α2(V) chains, encoded by the COL5A1 and COL5A2 genes respectively (Wenstrup et.al 2004). Mutations in the COL5A1 have been related to Ehlers-Danlos syndrome, which is characterized by reduced synthesis of collagen type V, causing irregularly large collagen fibrils within connective tissue (Malfait et.al. 2005).

One of the common single nucleotide polymorphisms of COL5A1, rs12722 C/T, located in the functional 3′ untranslated region (3′ UTR) on chromosome 9, is associated with tendon and ligament injuries. In particular, the CC genotype of rs12722 was found in higher proportion among asymptomatic control participants (participants without tendinopathy) in South Africa (Mokone et al. 2006) and in Australia (September et al. 2008). Therefore, it was concluded that the CC genotype of rs12722 is associated with reduced risk of chronic Achilles tendinopathy. The meta-analysis done by Pabalan et.al. concluded that Col5A1 polymorphisms reduce the risk of tendon-ligament injuries (TLI) among Caucasians. In addition, the rs12722 was associated with anterior cruciate ligament injury (ACL) in females (Posthumus et.al.2009), joint range of motion (Brown et al. 2011), lateral epicondylitis (Altinisik et.al. 2009) and exercise-associated muscle cramping (EAMC) (O’Connell et.al. 2013). The TT genotype of the COL5A1 rs12722 gene variant has been associated with enhanced endurance running performance (Posthumus et al. 2011; Brown et al. 2011a; 33O’Connell, Posthumus & Collins 2014). It has been suggested that the TT allele at the rs12722 variant results in increased type V collagen production, altering the mechanical properties of the tissue, leading to improved endurance performance. Based on this, it was hypothesized that rs12722 could change the structural and mechanical properties of tendons and ligaments, such as degree of stiffness and thickening. Kirk et. al. supported this claim that TT genotype of rs12722 is associated with greater tendon stiffness, while Forster et.al. reported no association between tendon properties and rs12722 (Kirk et al., 2016; Foster et al., 2014).

The possible mechanism between COL5A1 gene variants and soft tissue injuries was proposed by Laguett et.al. The authors stated that the region where rs12722 is located (COL5A1 3′ UTR) affects mRNA stability (Abrahams et al.). The 3-untranslated region (UTR) of eukaryotic genes has important post-transcriptional regulators (Mazumder et al., 2003; Xie et al., 2005) and plays a significant role in disease etiology (Sayed & Abdellatif, 2011). Altered COL5A1 mRNA stability has been proposed to result in altered type V collagen production, changes in collagen fibril diameter and packing density, and potentially altering the mechanical properties of connective tissues (Collins et al., 2011). Thus different genotype composition of rs12722 might change the co-polymerisation between collagen type V and type I fibrils (Abrahans et al.,; Llaguette et al., 2011). However, it has not been demonstrated experimentally and exact mechanisms are still unknown. It has been reported that the T alleles of rs12722 is associated with increased mRNA stability, resulting in increased α1(V) chain and type V collagen production, decreasing fibril diameter and packing density, and altering the mechanical properties of tendons and other connective tissues. The mRNA stability of the CC genotype was significantly lower compared to the TT genotype, suggesting that less α1 chain of type V collagen is synthesized from the C-form (Laguette et al., 2011).

Both collagen type I and type V can affect the proliferation of TB infection since both of them constitute the lung parenchyma, the main disease site. However, the contribution of polymorphisms of the collagen gene (Col1A1 and Col5A1) and metalloproteinases (MMP) to human TB susceptibility remains poorly studied. The following population-based preliminary case-control study of adults with TB disease and controls without TB disease will evaluate associations between rs1800012 in COL1A1, rs1272222 in COL5A1 and rs679620 in MMP3 genes human TB susceptibility in Kazakhstan.

## METHODS

### Participants

162 samples were obtained for the study. Demographic information, genetic data and environmental determinants of the TB susceptibility from the Kyzylorda, Kostanay, Almaty oblast and Almaty city regions of the Kazakhstan Republic between 2012 and 2014 were analsyed. Participants were organized into two different groups representing 112 TB-free controls and 50 cases with pulmonary TB. Patients were considered to be pulmonary tuberculosis cases according the positive *Mycobacterium Tuberculosis* culture, smear microscopy or on clinical and radiological data (Guidelines on Tuberculosis Control in the Republic of Kazakhstan, 2008). Controls are those that do not have prior pulmonary tuberculosis diagnosis. Inclusion criteria included the requirement that participants must be at least 18 years of age at the time of screening, having a permanent living place in which the participants have lived for at least 3 months and no previous history of serious mental impairments which could invalidate informed consent provision or further participation in the study.

### Data collection

All the study participants completed the informed consent and a questionnaire specifying age, gender, socioeconomic status. Blood samples were collected from the participants who agreed to further participation in the research.

### SNP analysis of samples

Samples were collected into 2 microcentrifuge Eppendorf tubes and labelled with a unique ID for each participant. The centrifugation was performed in order to separate the sample into components of plasma and serum. All the blood components were kept in special containers in the refrigerator with −80°C temperature regimen. Blood DNA extraction was carried out using Wizard Genomic DNA Purification Kit (Promega, Madison, Wisconsin, USA) following manufacturer’s protocol. The concentration and purity of the extracted DNA were measured with NanoDrop spectrophotometer. SNPs were genotyped on an SNP array using qualitative real-time

PCR on a Life Technologies Quantstudio 12K Real Time PCR system using the cycle relative threshold (Crt) method. The results were entered into the Life Technologies Copy Caller software and examined. A two assay reaction with a pair of primers and Taqman probe were used for each single nucleotide polymorphism target, they are rs1800012, rs1272222.

### Ethics

Research protocols were reviewed and approved by the National Laboratory Astana Ethics Committee Center of Life Sciences.

## Statistical analysis

The STATA version 14.2 for Windows was used to perform univariate descriptive, bivariate X^2^ and multiple logistic regression analysis. The Hardy-Weinberg equilibrium test and bivariate X ^2^ statistics for the different inheritance patterns were made using the SNPStats web tool. Demographic and personal characteristic variables were collected and categorized as needed utilizing STATA 14.2. All variables were coded as dichotomous dummy variables and frequencies were computed. In bivariate analysis, all variables were analyzed using simple logistic regression with the outcome variable TB disease status (TB diseases versus no TB disease), producing unadjusted odds ratios (ORs), 95% CIs and p-values. The candidate SNPs were analyzed for independence utilizing the Hardy-Weinberg equilibrium (HWE). The associations between SNPs and the TB disease status outcome were evaluated using unadjusted and adjusted ORs, 95% CIs and p-values, computed from bivariate logistic regression and multivariate logisitic regression models respectively. If a variable showed a p-value<0.25 in bivariate analysis, it was included in multivariate logistic regression modeling as a covariate for final analysis. Covariates were retained in the final multivariate logistic regression model if their associated p-value was less than or equal to .05, considered statistically significant. Interactions between covariates were tested for the outcome TB disease status but no statistically significant interactions at the α=.05 level were identified.

## Results

Distributions between cases and controls for almost all demographic and personal characteristic variables are adequate for further analyses (Table 1). Table 2 shows that the categorical variables Age, BMI, Job status and Alcohol consumption and HIV status were statistically significantly (p-value<0.05) associated with TB infection. As the borderline p-value≤0.25 was set up as a criterion for including into the finalized multiple logistic regression model, the variable Location Type (p=0.148) was also considered (Table 2). Variables such as Sex, Ethnicity and Risk Factor Smoking were found to be insignificant and did not vary between case and control groups (Table 2).

**Table 1.**
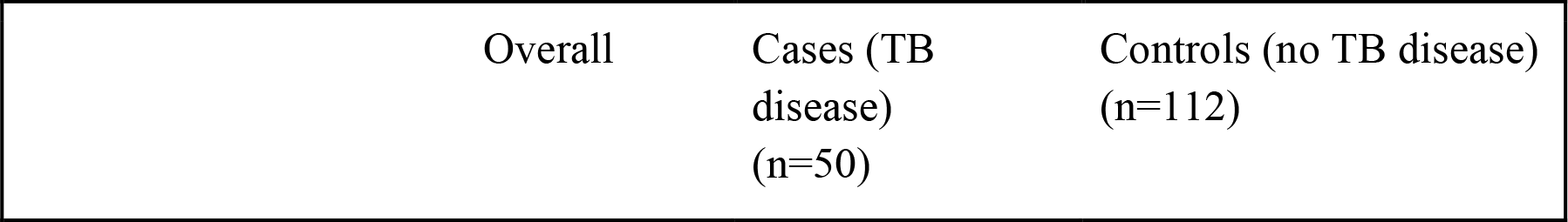
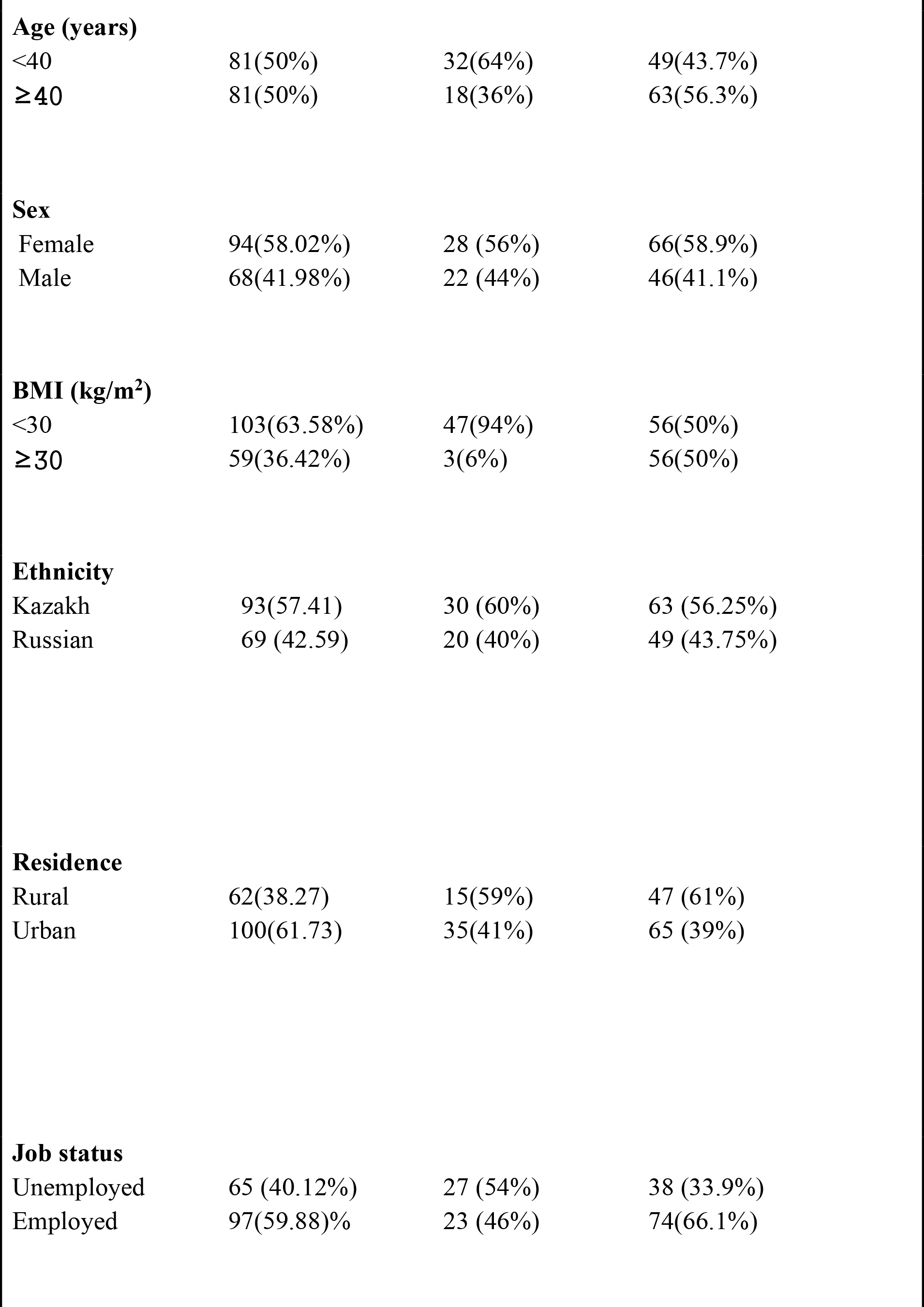
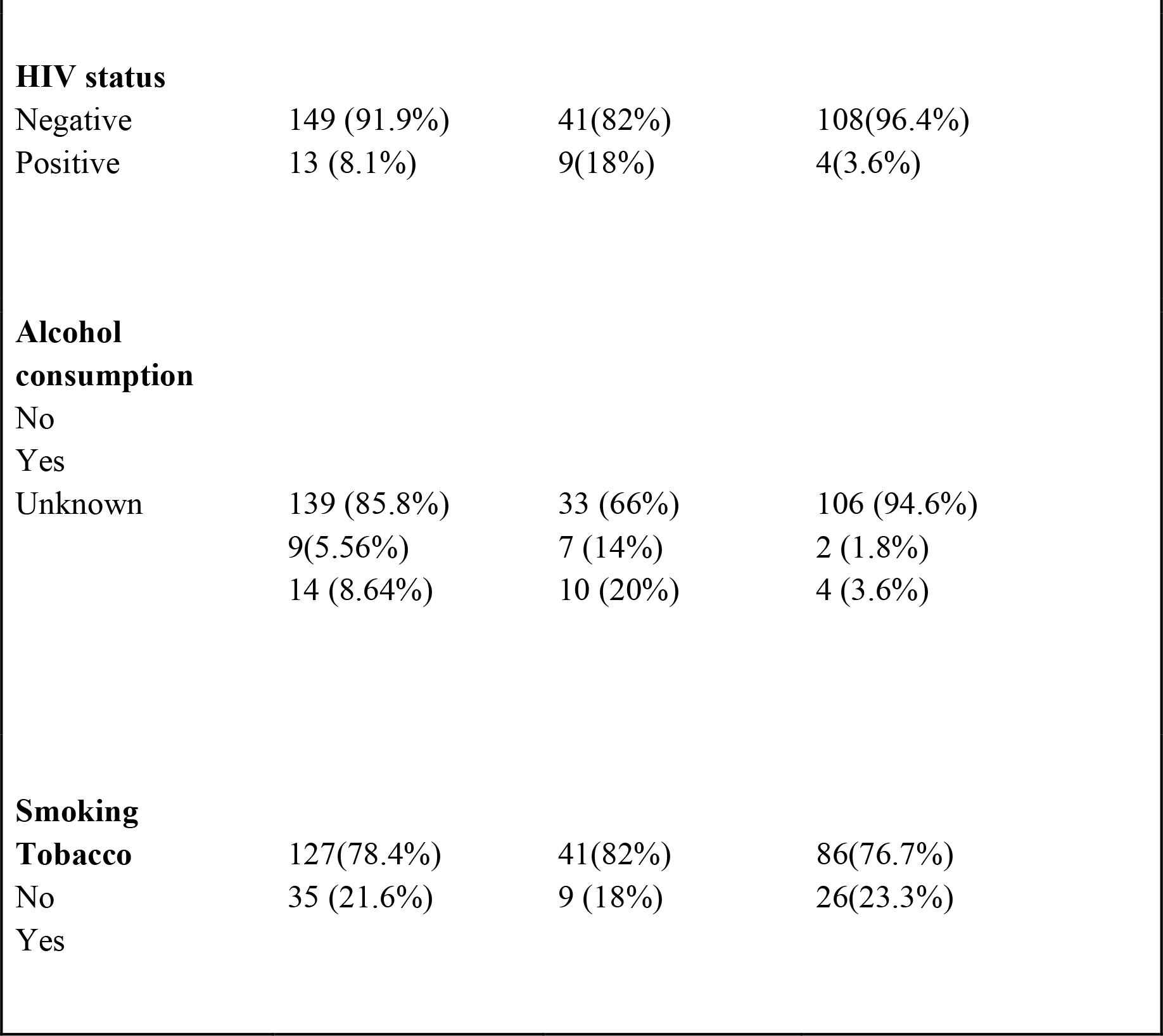
Descriptives of demographic and personal characteristics by case-control status.

| Overall | Cases (TB disease)<br>(n=50) | Controls (no TB disease)<br>(n=112) |
| --- | --- | --- |

|  |  |  |  |
| --- | --- | --- | --- |
| **Age (years)** |  |  |  |
| <40 | 81(50%) | 32(64%) | 49(43.7%) |
| ≥40 | 81(50%) | 18(36%) | 63(56.3%) |
| **Sex** |  |  |  |
| Female | 94(58.02%) | 28 (56%) | 66(58.9%) |
| Male | 68(41.98%) | 22 (44%) | 46(41.1%) |
| **BMI (kg/m<sup>2</sup>)** |  |  |  |
| <30 | 103(63.58%) | 47(94%) | 56(50%) |
| ≥30 | 59(36.42%) | 3(6%) | 56(50%) |
| **Ethnicity** |  |  |  |
| Kazakh | 93(57.41) | 30 (60%) | 63 (56.25%) |
| Russian | 69 (42.59) | 20 (40%) | 49 (43.75%) |
| **Residence** |  |  |  |
| Rural | 62(38.27) | 15(59%) | 47 (61%) |
| Urban | 100(61.73) | 35(41%) | 65 (39%) |
| **Job status** |  |  |  |
| Unemployed | 65 (40.12%) | 27 (54%) | 38 (33.9%) |
| Employed | 97(59.88)% | 23 (46%) | 74(66.1%) |

|  |  |  |  |
| --- | --- | --- | --- |
| **HIV status** |  |  |  |
| Negative | 149 (91.9%) | 41(82%) | 108(96.4%) |
| Positive | 13 (8.1%) | 9(18%) | 4(3.6%) |
| **Alcohol consumption** |  |  |  |
| No |  |  |  |
| Yes |  |  |  |
| Unknown | 139 (85.8%) | 33 (66%) | 106 (94.6%) |
|  | 9(5.56%) | 7 (14%) | 2 (1.8%) |
|  | 14 (8.64%) | 10 (20%) | 4 (3.6%) |
| **Smoking Tobacco** |  |  |  |
| No | 127(78.4%) | 41(82%) | 86(76.7%) |
| Yes | 35 (21.6%) | 9 (18%) | 26(23.3%) |

**Table 2.** Unadjusted bivariate association test between demographic and personal characteristic variables with the outcome TB disease status.

| Variable | Unadjusted OR | 95% CI | p-value |
| --- | --- | --- | --- |
| Age | 0.4375 | 0.22 - 0.87 | 0.017* |
| BMI | 0.064 | 0.0188 - 0.217 | <0.005** |
| Sex | 1.127 | 0.5749-2.211 | 0.727 |
| Ethnicity | 0.857 | 0.435 - 1.69 | 0.656 |
| Location type | 1.687 | 0.83 - 3.44 | 0.148 |
| Social status | 0.437 | 0.222 - 0.863 | 0.016* |
| Risk factor HIV | 5.926 | 1.73 - 20.31 | 0.004** |
| Risk factor Alcohol consumption | 3.446 | 1.846 - 6.434 | <0.005** |
| Risk factor Smoking | 0.726 | 0.312 - 1.69 | 0.456 |

\* - statistically significant with p-value<0.05 and $\geq 0.01$
\*\* highly statistically significant with p-value<0.01
†p-value<0.25 was set as the cutoff to be included for testing in the final multivariate logistic regression model with adjustments

### The candidate SNPs association with TB susceptibility

The COL1A1 and COL5A1 candidate SNPs rs1800012 and rs12722 were analyzed by the allele frequencies and genotype frequencies (Table 3). There were no differences observed in allele frequencies and genotype frequencies of COL1A1 SNP rs1800012 between case and control groups (Figure 1, Table 4). It was found out that no heterogeneity in allele frequency distribution, but some difference in genotype frequencies of COL5A1 SNP rs12722 exists between cases and controls (Figure 2, Table 4). The HWE test of independence have shown that case and control groups for both SNPs rs12722 and rs1800012 were independent (p-value>0.05) and it was possible to perform further tests on associations (Table 5). The different inheritance patterns TB susceptibility unadjusted association of candidate SNPs were analyzed (Table 6a, Table 6b). For the SNP rs1800012, none of the inheritance patterns have shown the association with positive TB status (Table 6a). However, there were associations with TB susceptibility shown to be significant (Table 6b). The overdominant mode of inheritance is shown to be statistically significant (p=0.023) and the heterozygous patterns were 2.187 times more likely to increase susceptibility for tuberculosis (Table 6b).

**Table 3.** Candidate SNPs and their descriptions.

| rs number | Gene | Risk allele | Function |
| --- | --- | --- | --- |
| rs12722 | COL5A1 | T | Associated with Achilles tendinopathy and Ehlers-Danlos syndrome |
| rs1800012 | COL1A1 | T | This allele has association with abnormal bone marrow density and increased risk of intervertebral disc disease, osteoporosis and fractures. |

**Table 4.**
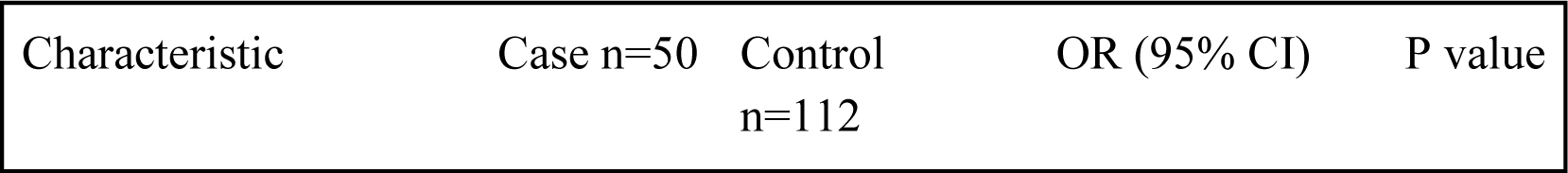
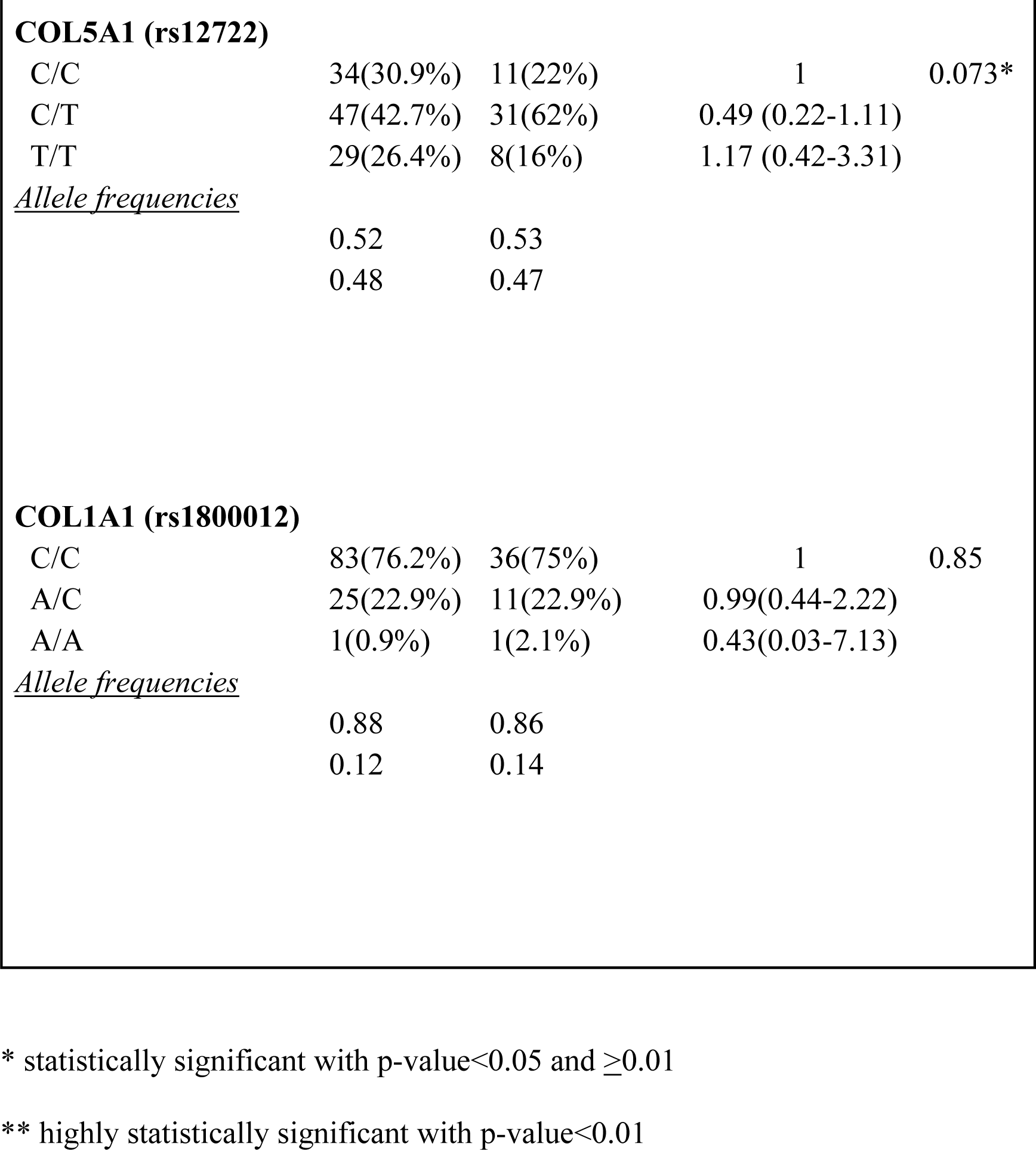
Allele and genotype distribution of two SNPs of COL5A1 and COL1A1 genes.

**Table 5.** Hardy-Weinberg equilibrium P values for collagen SNPs.

| SNP rs number | Hardy-Weinberg equilibrium p-value |  |
| --- | --- | --- |
|  | Cases(n=50) | Controls (n=112) |
| rs1800012 | 1 | 1 |
| rs12722 | 0.15 | 0.13 |

**Table 6a.**
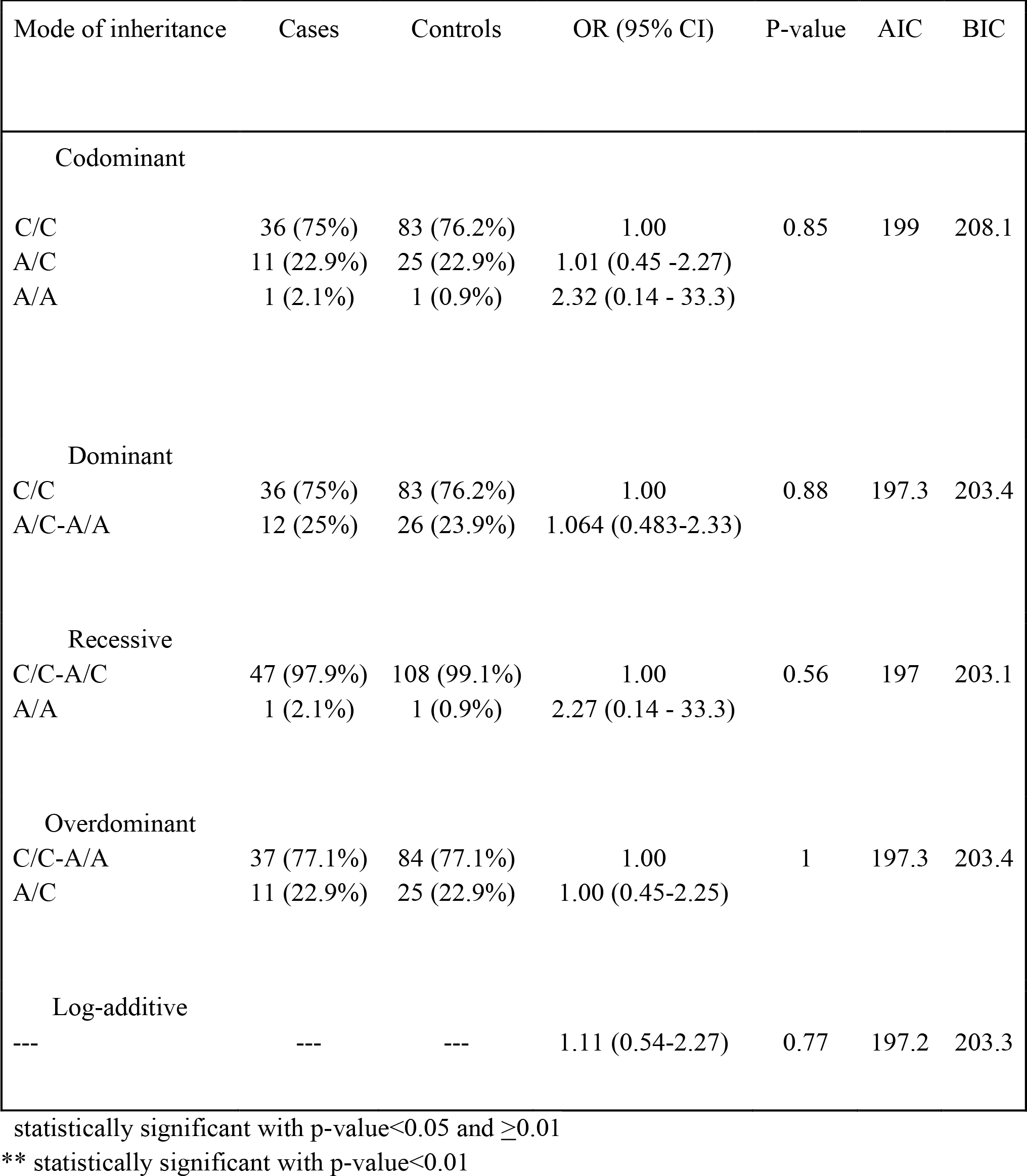
p-values for different inheritance patterns of rs1800012.

| Mode of inheritance | Cases | Controls | OR (95% CI) | P-value | AIC | BIC |
| --- | --- | --- | --- | --- | --- | --- |
| Codominant |  |  |  |  |  |  |
| C/C | 36 (75%) | 83 (76.2%) | 1.00 | 0.85 | 199 | 208.1 |
| A/C | 11 (22.9%) | 25 (22.9%) | 1.01 (0.45 -2.27) |  |  |  |
| A/A | 1 (2.1%) | 1 (0.9%) | 2.32 (0.14 - 33.3) |  |  |  |
| Dominant |  |  |  |  |  |  |
| C/C | 36 (75%) | 83 (76.2%) | 1.00 | 0.88 | 197.3 | 203.4 |
| A/C-A/A | 12 (25%) | 26 (23.9%) | 1.064 (0.483-2.33) |  |  |  |
| Recessive |  |  |  |  |  |  |
| C/C-A/C | 47 (97.9%) | 108 (99.1%) | 1.00 | 0.56 | 197 | 203.1 |
| A/A | 1 (2.1%) | 1 (0.9%) | 2.27 (0.14 - 33.3) |  |  |  |
| Overdominant |  |  |  |  |  |  |
| C/C-A/A | 37 (77.1%) | 84 (77.1%) | 1.00 | 1 | 197.3 | 203.4 |
| A/C | 11 (22.9%) | 25 (22.9%) | 1.00 (0.45-2.25) |  |  |  |
| Log-additive |  |  |  |  |  |  |
| --- | --- | --- | 1.11 (0.54-2.27) | 0.77 | 197.2 | 203.3 |

statistically significant with p-value<0.05 and $\geq 0.01$
\*\* statistically significant with p-value<0.01

**Table 6b.**
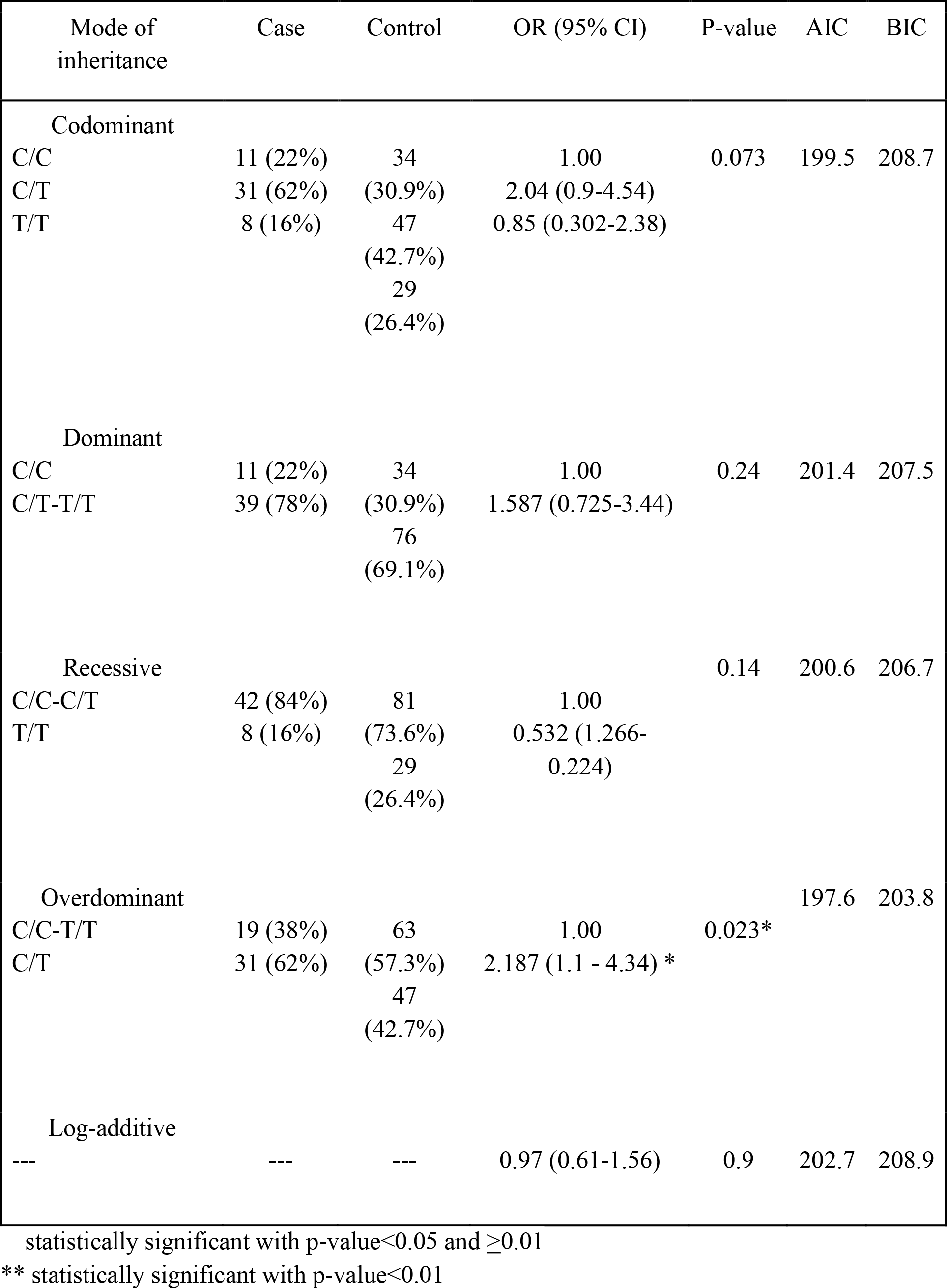
p-values for different inheritance patterns of rs12722.

**Figure 1:**
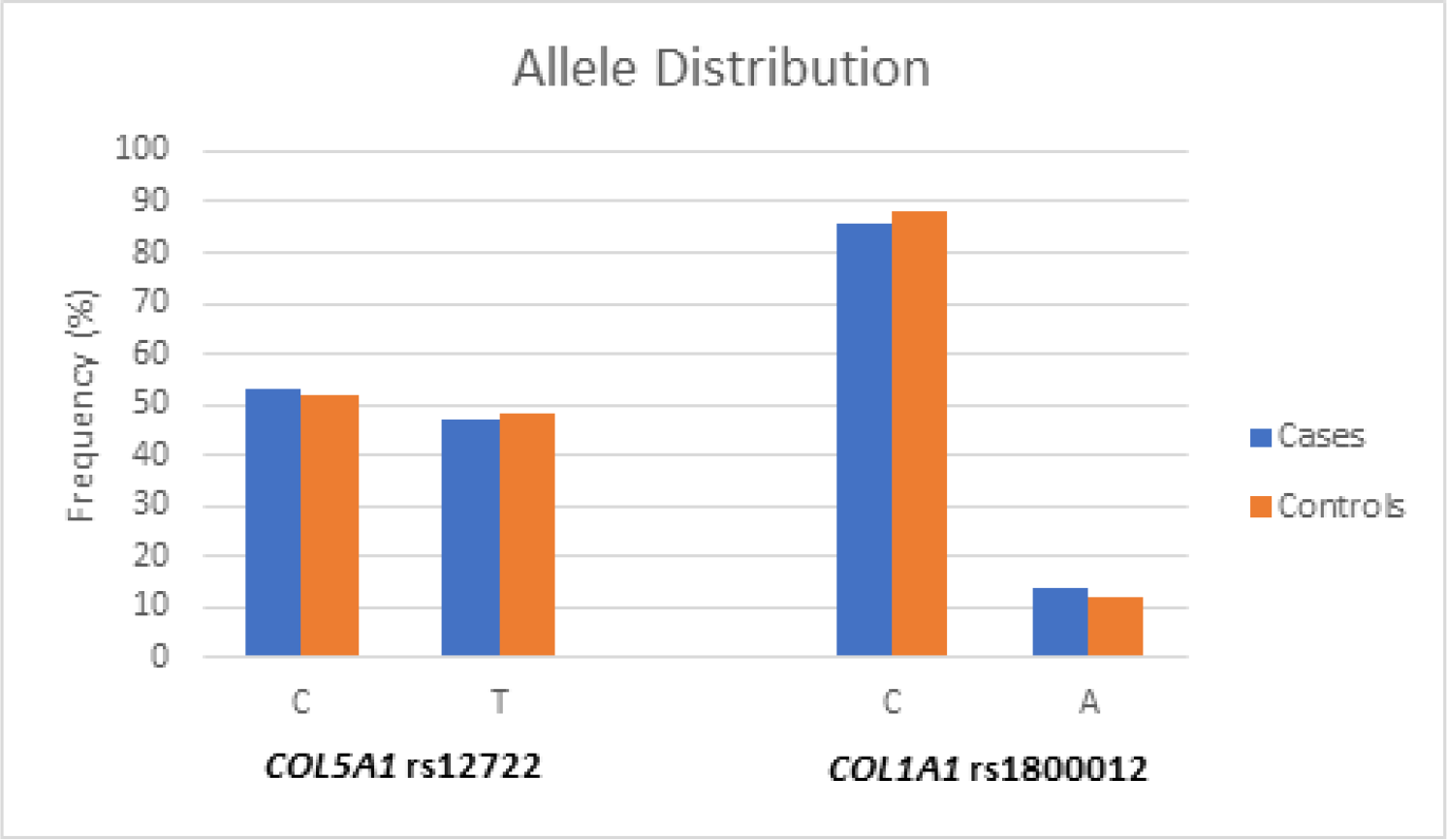
Allele frequencies of SNPs of COL5A1 and COL1A1 genes.

**Figure 2:**
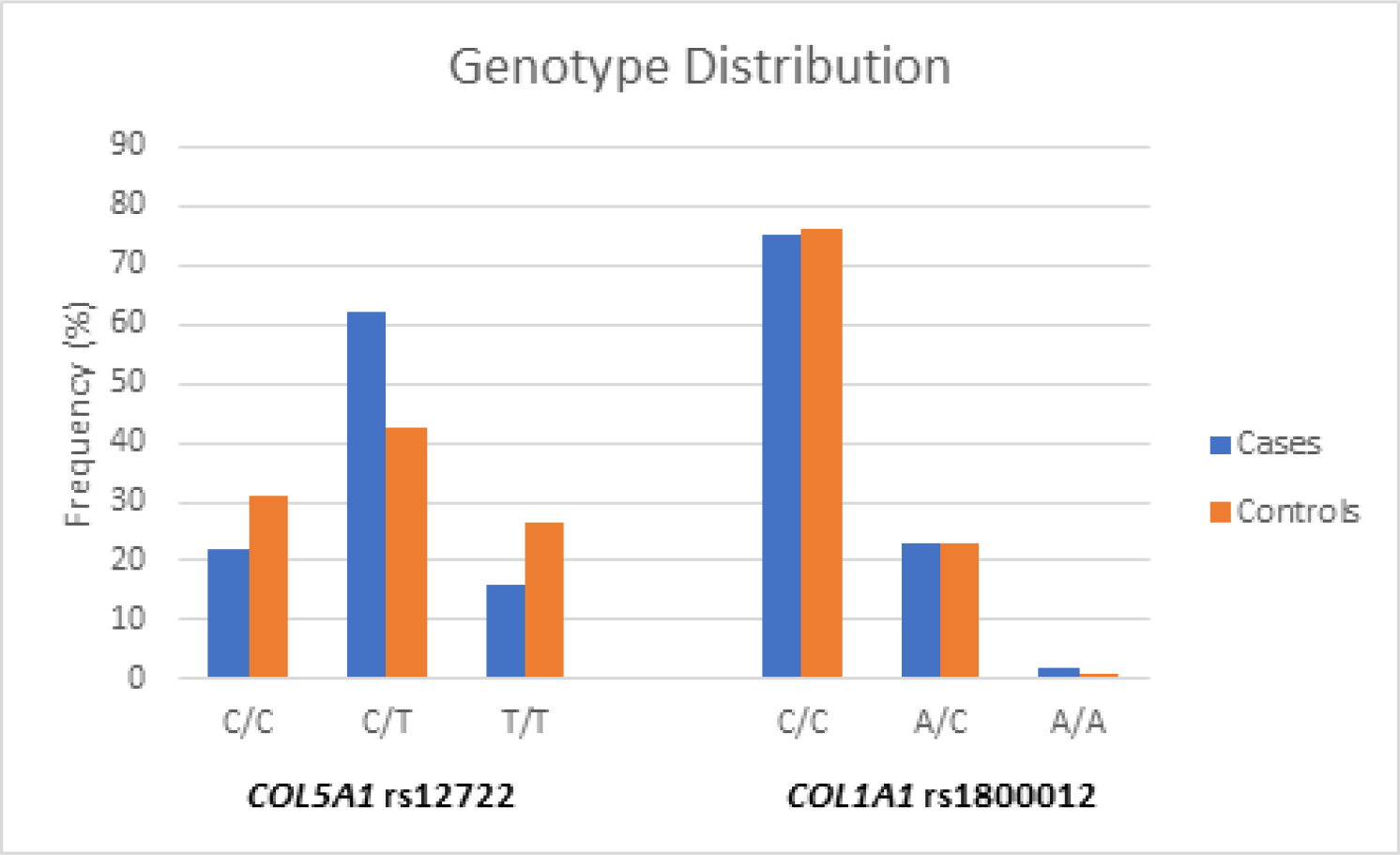
Genotype frequencies of SNPs of COL5A1 and COL1A1 genes.

### The multiple logistic regression analysis of TB status in relation with all the statistically significant demographic variables and SNPs

Multiple regression model included only statistically significant demographic variables and one statistically significant SNP rs12722 with an overdominant inheritance pattern as an explanatory variable (Table 7). The multicollinearity and interactions between predictors have been assessed before setting up the finalized model. The collinear patterns were observed between variables Age and BMI, Risk Factor Alcohol and Risk factor HIV. Predictors BMI and Risk Factor Alcohol were omitted from the final model. There was no effect modification observed between explanatory variables. The final model shows that Age was strongly associated with developing pulmonary TB (p-value=0.03) having odds of developing condition 2.27 times less for people older than 40 years of age adjusting for other variables in the model (Table 7). The Social status (p-value=0.02) is shown to increase the risk of developing TB for 2.38 times in unemployed people than employed adjusting for other variables. Being HIV-positive (p-value=0.004) has shown to significantly increase the risk of having TB for 6.9 times than in HIV-negative adjusting for other variables included in the final model. Having heterozygous allele of COL5A1 SNP rs12722 (p-value=0.019) is shown to increase the odds of developing TB 2.6 times than in patients with homozygous patterns adjusting for Age, Social status, Risk Factor HIV (Table 7).

**Table 7.**
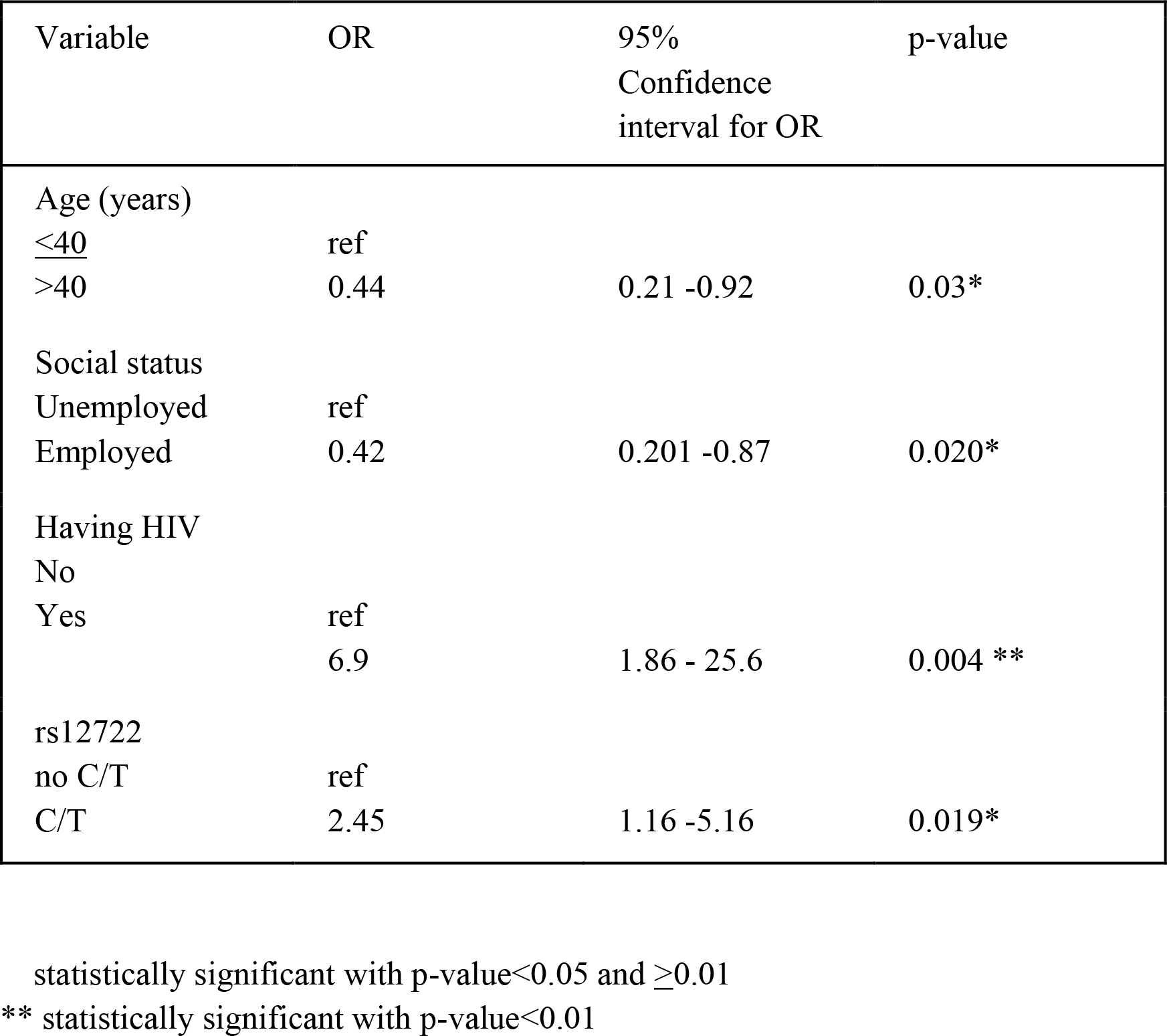
Final multivariate logistic regression model, including only statistically significant covariates with TB disease as the outcome.

## Discussion

The main finding of our study was that COL5A1 rs12722 polymorphisms are overrepresented in TB infected patients when compared to the control group. No previous studies have found the same results. The present study provides novel findings that contributes to the understanding of the role of COL5A1 gene polymorphisms in patients that are susceptible to TB. The previous studies evaluating rs12722 polymorphisms in COL5A1 gene mainly focus on ligaments and tendons because collagen comprises approximately 75% of their weight (Frank, 2004). Collagen is the primary component of extracellular matrix and connective tissues. and many diseases are caused by mutations in collagen genes or collagen degrading enzymes (Myllyharju, 2004). The SNP rs12722 is associated with a high risk of musculoskeletal soft tissue injuries such as tendon or ligament injuries (Lv et al., 2018). The most common condition that results from mutations of the COL5A1 gene is classic Ehlers-Danlos syndrome (Wenstrup et al., 2000). Type V collagen binds to other collagens to produce heterogeneous trimeric molecules that result in collagen scaffolding (Kirk et al., 2016). Therefore, it also plays an important role in regulating fibril diameter and density of the extracellular matrix of lung tissues. Since collagen builds the connective tissue fibers, the arrangement of lung structure is maintained mainly by collagen fibers (Al Shammari et al., 2015). Pulmonary cavitation is an important hallmark of TB pathogenesis Cavitation plays an important role in the leaking of tubercle bacilli into bronchial cavities. This cavitation depends on MMPs that are involved in ECM destruction (Mankaet al., 2012). Therefore, our findings support the hypothesis that there is an association between collagen degradation and immunopathology in TB.

Our findings also show that in COL5A1 (rs12722) polymorphism, the CT genotype is significantly associated with a high risk of TB disease. However, there was no significant difference in allele frequencies between TB-infected patients and TB-free control group. Different studies that have found associations between rs12122 (BstUI RFLP, C/T) and Achilles tendinopathy (September et al., 2009), anterior cruciate ligament ruptures (Posthumus, 2009) and endurance running performance (Brown et al., 2011). The CC genotype of rs12722 decreases the risk of developing Achilles tendinopathy, as compared to T allele (TT or TC genotypes) (September et al., 2009). In these studies, TT is associated with connective tissue stiffness. Stiffness is defined as a mechanical property of connective tissues to resist deformation (Foster et al. 2012). One of the characteristics of Ehlers-Danlos syndrome is a hypermobile joint that results from reduced stiffness (Eric et al., 2016). Therefore, lowered tendon stiffness lead to increased risk of connective tissue injuries.

Ourfindings show that COL1A1 rs1800012 polymorphism had no statistically significant association with TB-infected patients. The findings of our study do not support the hypothesis for an association between COL1A1 and TB pathogenesis. Type I collagen is one of the essential and most abundant components of connective tissues (Sequeglia et al., 2018). In other studies, it has been reported that rs1800012 in COL1A gene is associated with bone marrow density (Rojano et al., 2013, Zintzaras et al., 2011) and musculoskeletal degenerative diseases (Zhong et al., 2011), such as intervertebral disc degeneration and osteoarthritis.

The findings of the final logistic regression model show that the development of TB is also associated with age, employment, and HIV. The odds of having TB infection is statistically significantly higher among those who are old than those who are young. Disease risk depends on age-related differences accompanied by various clinical features and response to infection. According to the WHO, adults in their most productive years are the most affected group (WHO, 2018). The risk of infection reaches a peak in adults between aged 20-30 years because adults in this age have post-primary tuberculosis characterized by lung cavitation and tissue destruction

(Marais et al., 2004). However, the elderly population is also highly susceptibility to TB disease (Gardner, 1980). Globally, the highest rate of TB disease incidence is among people aged 40-45years, but in some countries in South East Asia, the Eastern Mediterranean and the Pacific Region TB disease rates peak in ages equal to or above65 years-of-age (WHO, 2014). Elderly people suffer greater TB mortality rates than the younger populations because aging leads to age-related decline in the functioning of the immune system (Stervbo et al., 2015).

Another study finding indicates that employment is protective against developing TB disease when compared to persons unemployed. There are several studies on a socioeconomic status that was conducted in countries with a high prevalence of TB disease finding that risk of TB increases with lower income (Hiransuthikul et al., 2004, Mahomed et al., 2011). Since unemployment is a marker of lower socioeconomic status and marginalization, it was expected that unemployment would be associated with a higher risk of TB infection—consitstent with our findings.

The findings also indicate that patients with HIV infection have a higher risk of being infected with TB as compared to those who do not have HIV infection, consistent with the literature (Narasimhan et al., 2013). Studies conducted in countries with high HIV burden have shown that prevalence of TB disease is strongly associated with the incidence of HIV infection (Corbett et al., 2003). Coinfection with HIV significantly increases the probability of reactivation of latent TB infection (Lawn, 2005). The reactivation of TB occurs due to weakened cell-mediated immunity that results from HIV infection.

Various anti-TB drugs have been used for the treatment of latent TB infection (Denholm and McBryde, 2010) to reduce the likelihood of the develop of TB disease. To prevent the reactivation of latent TB infection, isoniazid and rifampin have been utilized. Isoniazid prevents the spread of M. tuberculosis from granuloma by restricting the chemicals of tubercle bacilli that invade the host immune system (Takayama et al., 2005). Mycolic acid protects M. tuberculosis from immune cells-isoniazid inhibits mycolic acid to prevent the replication of tubercle bacilli (Denholm and McBryde, 2010). Enzymes that lead to collagen destruction are also important to limit the active TB infection. The MMP inhibitor Marimastat was shown to be effective to reduce the bacterial load and granuloma formation in a TB infected tissue model (Parasa et al., 2017). These MMP inhibitors also increase the effectiveness of isoniazid and rifampin (Xu et al., 2018). The USA has only one licensed MMP inhibitor, doxycycline, that is utilized to suppress the effect of collagenase activity (Gapski et al., 2009). Clinical trials have demonstrated that doxycycline reduces immune-mediated tissue damage in different inflammatory conditions such as arthritis (Hanemaaijer et al., 1997), Gram-positive infections (Damian, 2006) and atherosclerosis (Lindeman et al., 2009). Thus, different studies show the importance of treatments that include a combination of MMP inhibitors and isoniazid as a strategy to increase drug efficacy. Given the current findings, further studies should focus on collagenase inhibitors to improve their specificity and efficacy.

## Data Availability

Request for data can be made to

## Acknowledgements

This work is in part supported by the Nazarbayev University School of Medicine Social Policy Grant 2016. The authors declare that they have no conflict of interest. CCK, SZ, AyK ArK conceptualized, design and coordinated the investigation. CCK, SZ, AyK ArK, AI, BC analyzed the data and carried out the statistical analysis. DE, AAk, UK participated in sample collection and DNA extraction. All authors participated in writing, reading and approval of the final manuscript.

We are grateful to Columbia University Global Health Research Center of Central Asia, Almaty and National TB Center, Almaty for organizational support of study participant recruitment. This work was partially supported by the Ministry of Education and Science of the Republic of Kazakhstan (within program # 0111RK00442)

